# A novel art of continuous non-invasive blood pressure measurement

**DOI:** 10.1101/2019.12.10.19013938

**Authors:** Jürgen Fortin, Dorothea Rogge, Christian Fellner, Doris Flotzinger, Julian Grond, Katja Lerche, Bernd Saugel

## Abstract

Wearable sensors to continuously measure blood pressure (BP) and derived cardiovascular variables have the potential to revolutionize patient monitoring. Current wearable methods analyzing time components (e.g., pulse transit time) still lack clinical accuracy, whereas existing technologies for direct BP measurement are too bulky. Here we present a new art of continuous non-invasive arterial blood pressure monitoring (CNAP2GO). It directly measures BP by using a new “volume control technique” and could be used for small wearable sensors integrated in a finger ring. As a software prototype, CNAP2GO showed excellent BP measurement performance in comparison with invasive BP in 46 patients having surgery. The resulting pulsatile BP signal carries information to derive cardiac output and other hemodynamic variables. We show that CNAP2GO can be miniaturized for wearable approaches. CNAP2GO potentially constitutes the breakthrough for wearable sensors for blood pressure and flow monitoring in both ambulatory and in-hospital clinical settings.

## Introduction

The digital revolution is about to bring major innovations to physiologic monitoring [^1^]. Innovative wearable sensors for the monitoring of cardiovascular dynamics have the potential to revolutionize the monitoring of vital signs and their response to therapy in both ambulatory and in-hospital clinical settings. Continuous real-time monitoring of cardiovascular dynamics – including arterial blood pressure (BP) and advanced hemodynamic variables that are derived from its waveform (stroke volume, cardiac output (CO), and dynamic cardiac preload variables) – is a mainstay of patient care in perioperative and intensive care medicine but, for the most part, still requires invasive or stationary non-invasive sensors [^2, 3^]. With the availability of non-invasive and wearable sensors, advanced cardiovascular monitoring may become part of patient surveillance on the normal ward [^4^] and even outside the hospital [^5^]. For these clinical applications, wearable sensors have to fulfill all regulatory and clinical demands of medical-grade devices – just as their stationary counterparts. Major challenges for future developments in the field of wearable miniaturized monitoring sensors will be to provide measurements with clinically acceptable accuracy, precision, and reliability and to ensure clinical usability and patient safety. At the moment, wearable sensors for continuous BP monitoring still show poor measurement performance [^6^] and have, therefore, not been adopted into clinical practice.

Most current wearable BP monitoring systems estimate BP based on time information. Pulse time intervals or pulse velocity methods use a proximal and a distal sensor for the measurement of the pulse transit time (PTT) or pulse arrival time (PAT) [^7^]. Other time-based methods for BP estimation use sensors that decompose the pulse into a forward and a backward wave to analyze their time differences [^8, 9^]. There are also attempts to use photo-plethysmographic (PPG) [^10^] or piezoelectric [^11^] pulse detection for the estimation of BP based on amplitude and time information. However, a direct translation of the pulse signal – which is a surrogate for volume changes – into continuous BP is challenging because of confounding effects of the cardiovascular, respiratory, and autonomic nervous systems. By enhancing model complexity (e.g., using machine learning and neural network tools), the measurement performance of these approaches may be improved [^10, 12, 13^]. Although their sensor hardware is very simple, significant work is still needed to achieve clinically acceptable performance [^7, 10, 14, 15^]. The difficulty with time-based approaches for wearable BP measurement devices seems to be that the variables of the mathematical models need to change along with changes in vasomotor activity of the vascular smooth muscles which are determined by the autonomic nervous system. Without a proper detection of changes in vasomotor activity, current models require frequent recalibration, for example using cuff-based sphygmomanometers, rendering them unfeasible for wearable wireless monitoring [^16^].

Besides time-based approaches, stationary finger-cuff devices have been successfully introduced for direct continuous non-invasive BP monitoring in various medical fields, especially in critical care and anesthesiology [^17^]. These devices are based on the “vascular unloading technique” (VUT), also referred to as “volume clamp” or “Peňáz” method [^18, 19, 20^]. The VUT directly measures BP and derives the complete pulsatile BP waveform by controlling fast inflation and deflation of a finger cuff. As with time-based technologies, the VUT is influenced by changes in vasomotor activity; different devices have found differing methods to detect and compensate for changes in vasomotor tone [^21, 22, 23^]. Existing systems are, however, too bulky to be used for wearable monitoring devices because the systems consist of mechanical elements like pumps, valves, and air hoses to follow the pulsatile nature of BP with high fidelity.

It was generally believed that, for correct BP measurement based on VUT, a constant (or “clamped”) blood volume in the finger over the complete period of pulsation and therefore full vascular unloading of the blood vessels is essential. Whether this is indeed physiologically or metrologically necessary was never examined.

The basic idea behind our novel art of continuous non-invasive BP measurement, “CNAP2GO”, is to perform the blood volume control substantially slower to be able to use slow-moving and, therefore, small-scaled hardware. Most importantly, such novel hardware requires neither a pump nor a valve. The basic principle of the CNAP2GO method is the *“volume control technique”* (VCT). In contrast to the VUT, which keeps the blood volume in the finger artery constant on a millisecond basis to accurately follow the full cardiac circle, the VCT keeps the volume constant on a time scale of cardiac cycles. Blood volume is allowed to oscillate over heart beats, balancing inflow and outflow of blood in the finger artery over each heart cycle. Controlling blood volume by VCT without fast oscillations poses a nontrivial control engineering problem, which is solved using a set of signal processing algorithms. Most importantly, as we will demonstrate, this method is resistant to changes in vasomotor tone.

In this paper, we first describe the elements of CNAP2GO and how they interact to derive a continuous BP waveform from the directly measured mean BP (mBP) and the PPG-signal. We will show that the BP waveform can be used to estimate advanced hemodynamic variables using pulse wave analysis. Currently, the VCT was implemented in software and placed on the hardware of an existing VUT-device that emulates slow VCT. The accuracy of the CNAP2GO software prototype was clinically validated in comparison with invasive BP measurements obtained using a radial arterial catheter in patients having surgery with general anesthesia. The CNAP2GO method can, in the future, be used for small wearable sensors integrated in a finger ring utilizing a light transmitter (LED) and a receiver (photodiode). The contact pressure of LED and photodiode can be modified by an actuator changing the ring diameter with a rate-of-change as fast (or rather: as slow) as mBP.

## Results

### Basic CNAP2GO considerations

The CNAP2GO sensor is a PPG-probe which comprises an LED and a photodiode. The contact pressure of these light elements to the skin can be modified. Even simple PPG-probes (e.g. pulsoxymeter) have elements that couple the light components to the body with pressure: this is usually a spring, Velcro, or another simple clamp mechanism. A constant pressure (around 20-30 mmHg) ensures good light coupling. Within CNAP2GO, a simple actuator can vary the contact pressure of the light elements as fast as mBP may change (Fig. 1). As a consequence, the system wins an additional degree of freedom which is needed for continuous BP measurement.

**Fig. 1:**
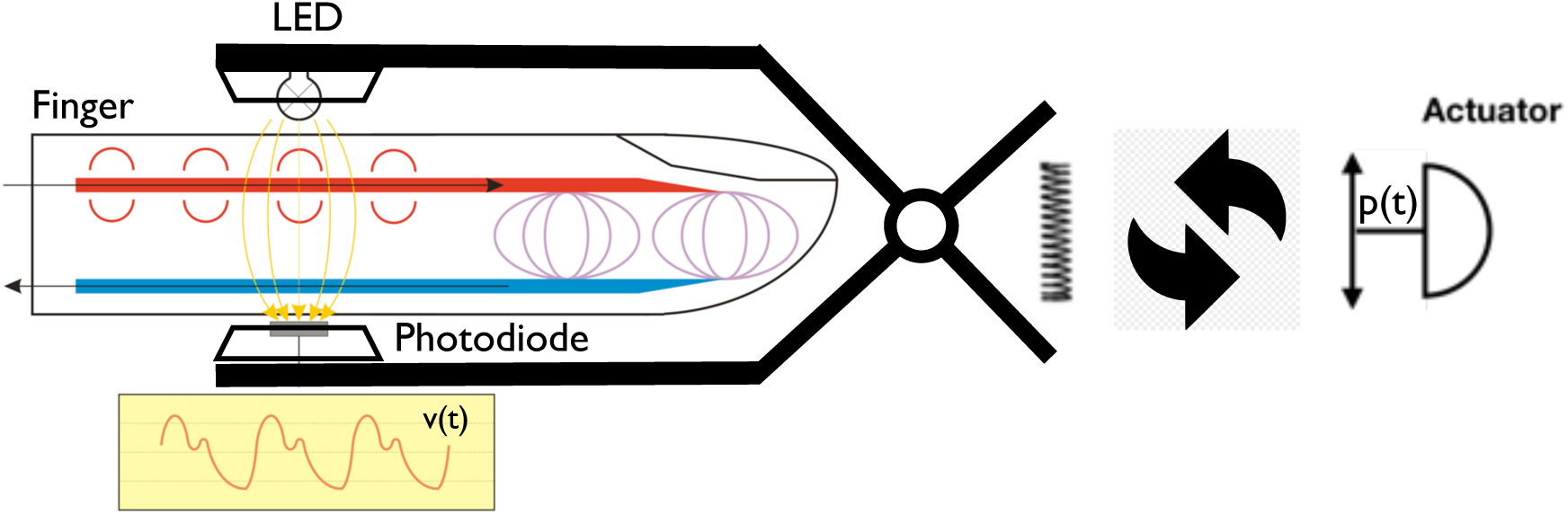
Within CNAP2GO, the element for constant pressure coupling is a simple, slow-moving actuator that can vary the contact pressure p(t) of the light elements as fast as mBP may change.

The basic idea of varying contact pressure is also implemented in all existing devices based on the VUT. When using the VUT, the speed and amplitude of the changes in contact pressure must match pulsatile arterial BP, requiring pressure changes of 30 – 100 mmHg per beat to ensure high-fidelity pressure waveforms. For example, the pressure system of the CNAP Monitor HD (CNSystems Medizintechnik, Graz, Austria) can apply pressure changes of up to 1500 mmHg/sec.

### CNAP2GO block diagram and signal flow

The CNAP2GO contact pressure is determined by a “closed-loop” control system using the PPG-signal and its components as input signals. Figure 2 shows the block diagram and the signal flow of CNAP2GO.

**Fig. 2:**
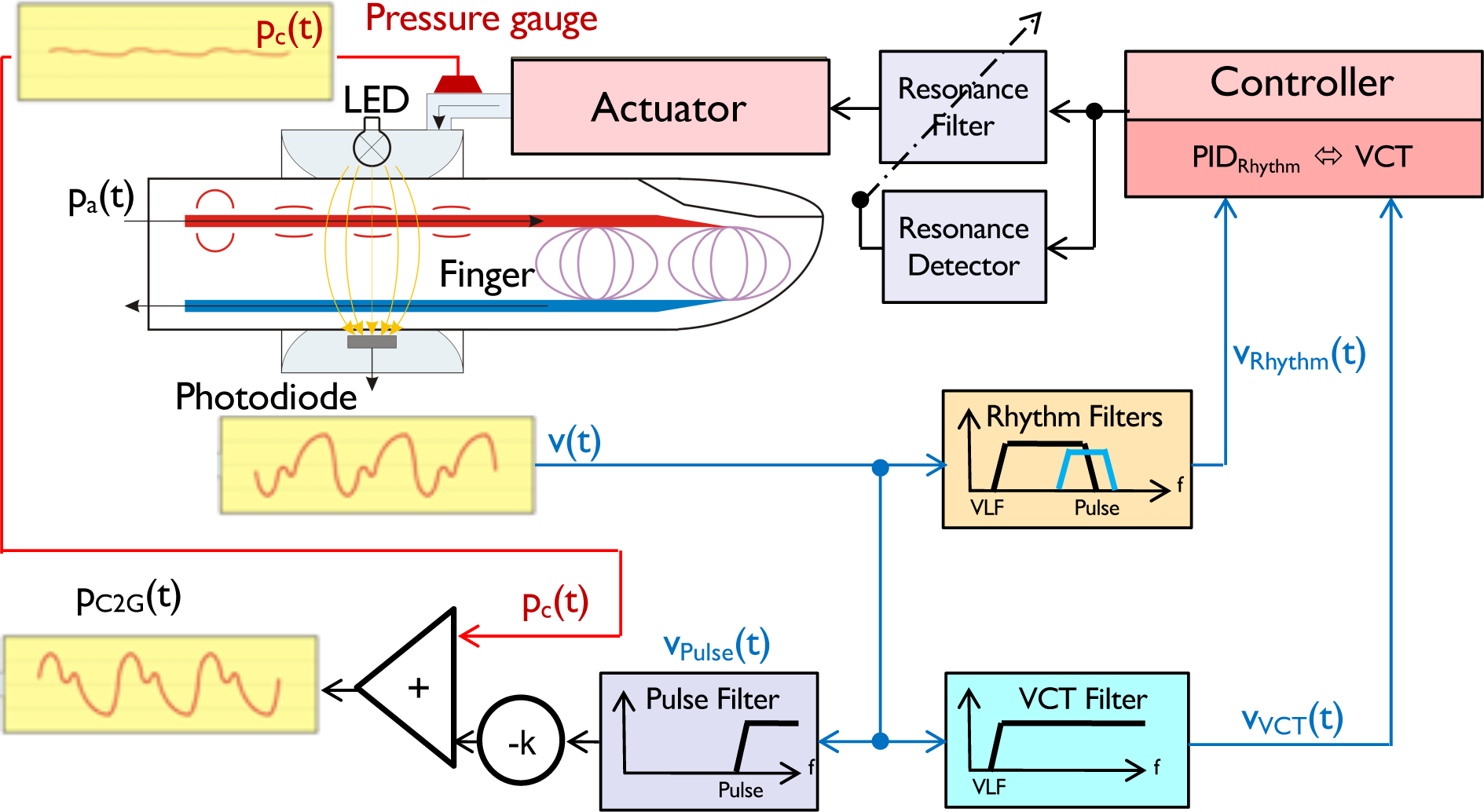
Block diagram and signal flow of CNAP2GO: An LED-light is directed through a finger and its absorption is measured by a photodiode. The resulting photo-plethysmographic signal v(t) is led into digital filters producing v_Pulse_(t), v_Rhythm_(t) and v_VCT_(t). Signal components v_Rhythm_(t) and v_VCT_(t) are used to control p_c_(t), whereas v_Pulse_(t) together with p_c_(t) are needed for the calculation of the pulsatile blood pressure signal p_C2G_(t). The control loop is closed using anti-resonance elements.

An LED-light (λ=890nm) is directed through a finger and light absorption is measured by a photodiode with corresponding wave-length characteristics. The resulting PPG signal v(t) is an inverse surrogate for blood volume changes inside the finger artery: Rising blood volume increases absorption and therefore decreases the amount of light transmitted through the finger.

The contact pressure p_c_(t) of the light elements can be altered by the actuator and is measured by a pressure gauge. v(t) is led into digital filters producing v_Pulse_(t), v_Rhythm_(t), and v_VCT_(t). The signal components v_Rhythm_(t) and v_VCT_(t) are used to control p_c_(t), whereas v_Pulse_(t) together with p_c_(t) are needed for the calculation of the pulsatile blood pressure signal p_C2G_(t). The control loop between the controller and the pressure is closed using anti-resonance elements. Note that v(t) and its components are dimensionless digital signals representing a surrogate of arterial volume in the finger.

### Finding the initial set-point

Closing the CNAP2GO control loop requires an initial open-loop phase to determine the initial value of mBP by altering the contact pressure and detecting the corresponding PPG pulse-amplitudes from the signal v_Pulse_(t). Figure 3 shows how a typical oscillometric curve including its envelope is obtained. Figure 3a shows the resulting v(t) when a pressure ramp p_c_(t) is applied. The p-v transfer function corresponds to an S-shaped arcus tangent with superimposed pulses [^24^]. The upper asymptote of v(t), where p_c_(t) is far above systolic BP (sBP), corresponds to the maximum light that can be detected from the finger when all blood is squeezed out and no pulses can be detected. The lower asymptote is at p_c_(t) = 0 where no deformation of the finger and its artery occurs (although this is a theoretical point of measurement because a minimum contact pressure is needed for proper light coupling). The amplitudes of the v(t)-pulses are derived from the high-pass filtered v_Pulse_(t). The amplitudes of v_Pulse_(t) are fitted using a Gaussian-style envelope curve, as can be seen in Figure 3b. The peak of this envelope indicates mBP according to the maximum oscillation rule [^25^], a principle which also holds true when using PPG signals [^26, 27, 28^]. After oscillometric envelope determination, the system applies mBP as initial contact pressure p_c_(t) and stores this value as the starting set-point p_0_ and its PPG-companion v_0_.

**Fig. 3:**
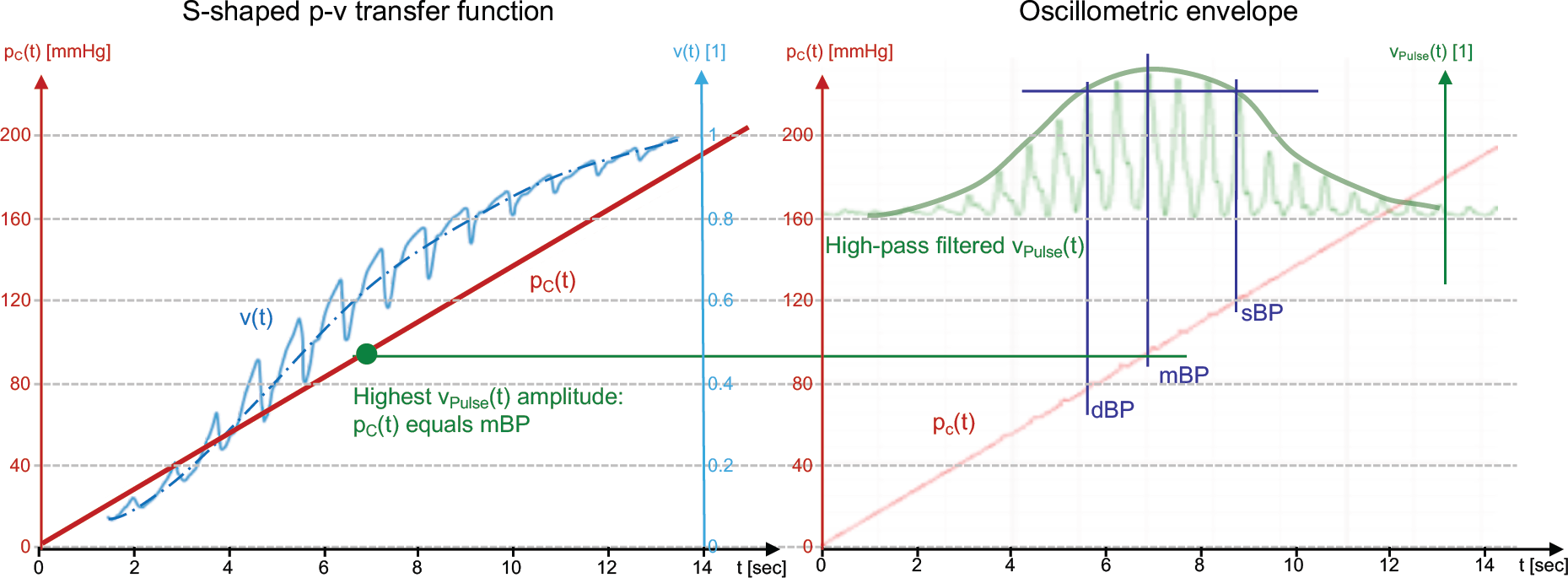
Depiction of a typical oscillometric curve including its envelope. (a) shows the resulting v(t) when a pressure ramp p_c_(t) is applied. (b) amplitudes of v_Pulse_(t) are fitted using a Gaussian-style envelope curve. See text for further explanation.

While this open-loop phase is very similar to the standard VUT, the closed loop of CNAP2GO differs in almost all fundamental aspects.

### Tracking mean BP

After the closed-loop phase, the mean cuff pressure is perfectly located on the inflection point of the S-shaped p-v transfer function. As can be seen in Figure 4a, the ideal pulsatile v_Pulse_(t) (light blue trace) has maximum amplitude and the integral of v_Pulse_(t) over a full beat is zero. Thus the main condition of the VCT, which keeps blood volume flow in the finger artery balanced over a heart cycle, is fulfilled.

**Fig. 4:**
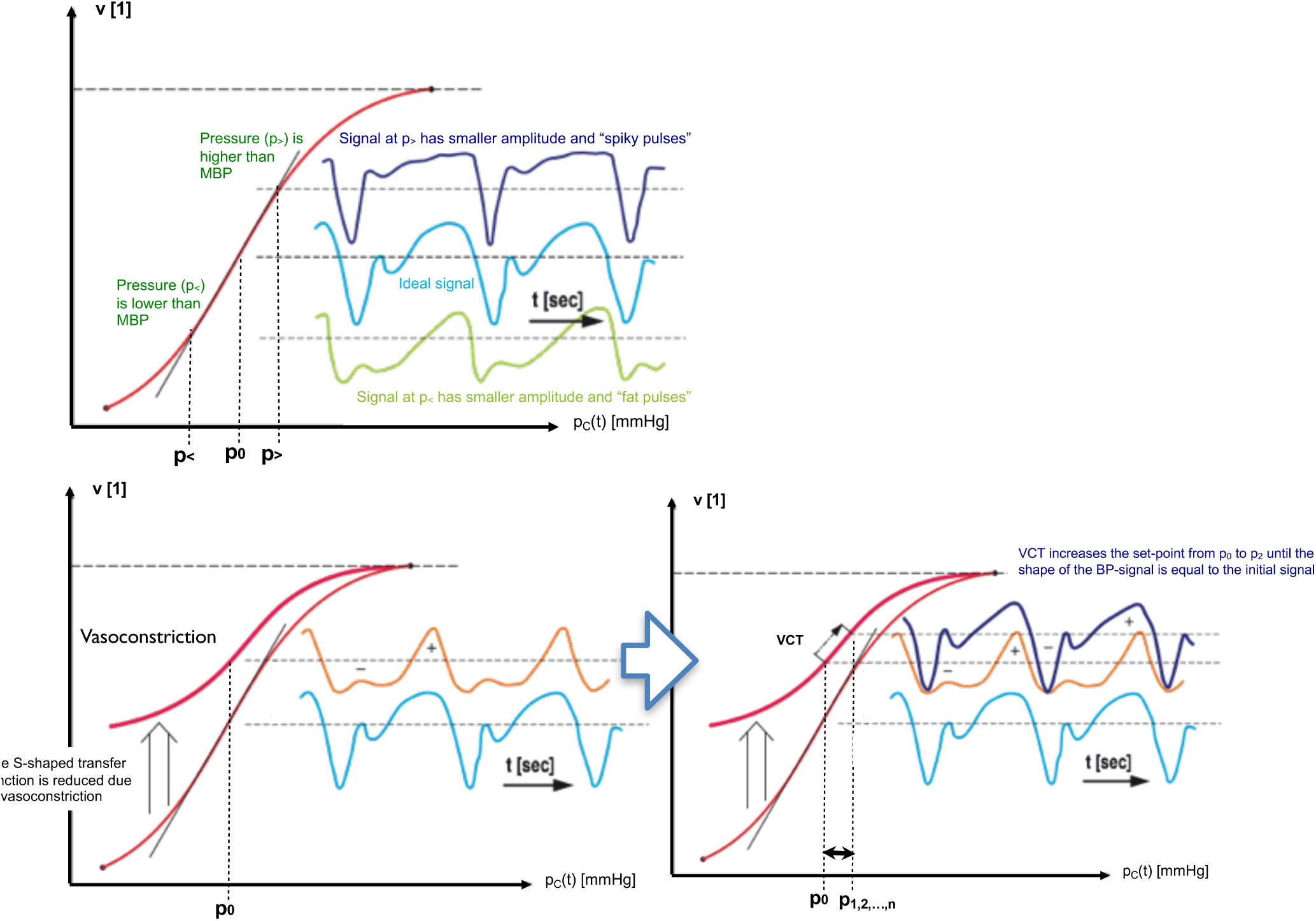
(a) The optimum pulsatile v_Pulse_(t) (light blue trace) has maximum amplitude with an integral over a full beat of zero. Suboptimal signals are either “spiky” (dark blue trace) or “fat” (green trace). (b) shows how vasoconstriction alters the S-shaped p-v transfer function. (c) shows how the set-point p_0_ and resulting p_C_(t) are altered after each heart beat. See text for further explanation.

As soon as BP or vasomotor changes occur, the situation becomes different. If the applied pressure is lower than mBP (p<), v_Pulse_(t) has a lower amplitude (green trace) and differs in pulse shape: the pulses are “fat”. In contrast, if the applied pressure is higher than mBP (p>), v_Pulse_(t) has also a lower amplitude, but the pulses are “spiky” (dark blue trace).

Figure 4b shows how vasoconstriction influences the p-v transfer function: while the upper asymptote for p_C_(t) >> sBP remains unchanged, the lower asymptote for p_C_(t) << diastolic BP (dBP) shows a higher v(t) because finger blood volume decreases during vasoconstriction. This results in a set-point lower than the inflection point and a change from v_0_(t) to a smaller but “fat” v_1_(t) with the negative half-wave of the “fat” pulsatile being greater than the positive half-wave. On the other hand, vasodilation increases the set-point over the inflection point, thus producing a “spiky” signal.

For both fat and spiky pulses, the integral of the pulsatile volume signal over a beat is no longer equal to zero. The CNAP2GO controller is responsible for restoring this condition of balanced volume control: In particular, the set-point p_0_ and resulting p_C_(t) need to be altered after each heart-beat until balance is achieved (Fig. 4c). A “proportional-integral” (PI) control approach results in an adaptive formula for set-point correction as can be seen in the methods section below. The summation of the “I” (integral) part of the control approach is the memory for the set-points p_1,2,…,n_ that allows for the reconstruction of long-term BP information.

This mechanism not only works for vasomotor changes but also for pure mBP changes. An increase in mBP – which is a shift of the p-v transfer function to the right – tends to move the setpoint towards the lower asymptote of the p-v transfer function. The integral over the beat will be negative resulting in an increase of the set-point p_n_. Vasodilation shifts the S-curve in the opposite way and VCT decreases p_n_.

Figure 5 shows the frequency domain of the CNAP2GO control signals. Beside the high-pass filtered pulses v_Pulse_(t), also the very-low-frequency components below 10^−2^ Hz of v(t) are removed from v_VCT_(t) and v_Rhythm_(t) by a special digital filter design. In these very slowly changing frequency components, blood volume and thus v(t) is not only influenced by BP but also by vasomotor activity. In the case of vasoconstriction, smooth vascular muscles close the arteries and arterioles and finger blood volume decreases (i.e., v(t) goes up), although BP typically rises during vasoconstriction (i.e., v(t) should go down). The opposite behavior occurs during vasodilation. This means that the information from the PPG signal v(t) below 10^−2^ Hz is not reliable. Therefore, this frequency range is removed from all further calculations by filtering. The above-mentioned “I” summation of the VCT control approach reconstructs BP information below 10^−2^ Hz.

**Fig. 5:**
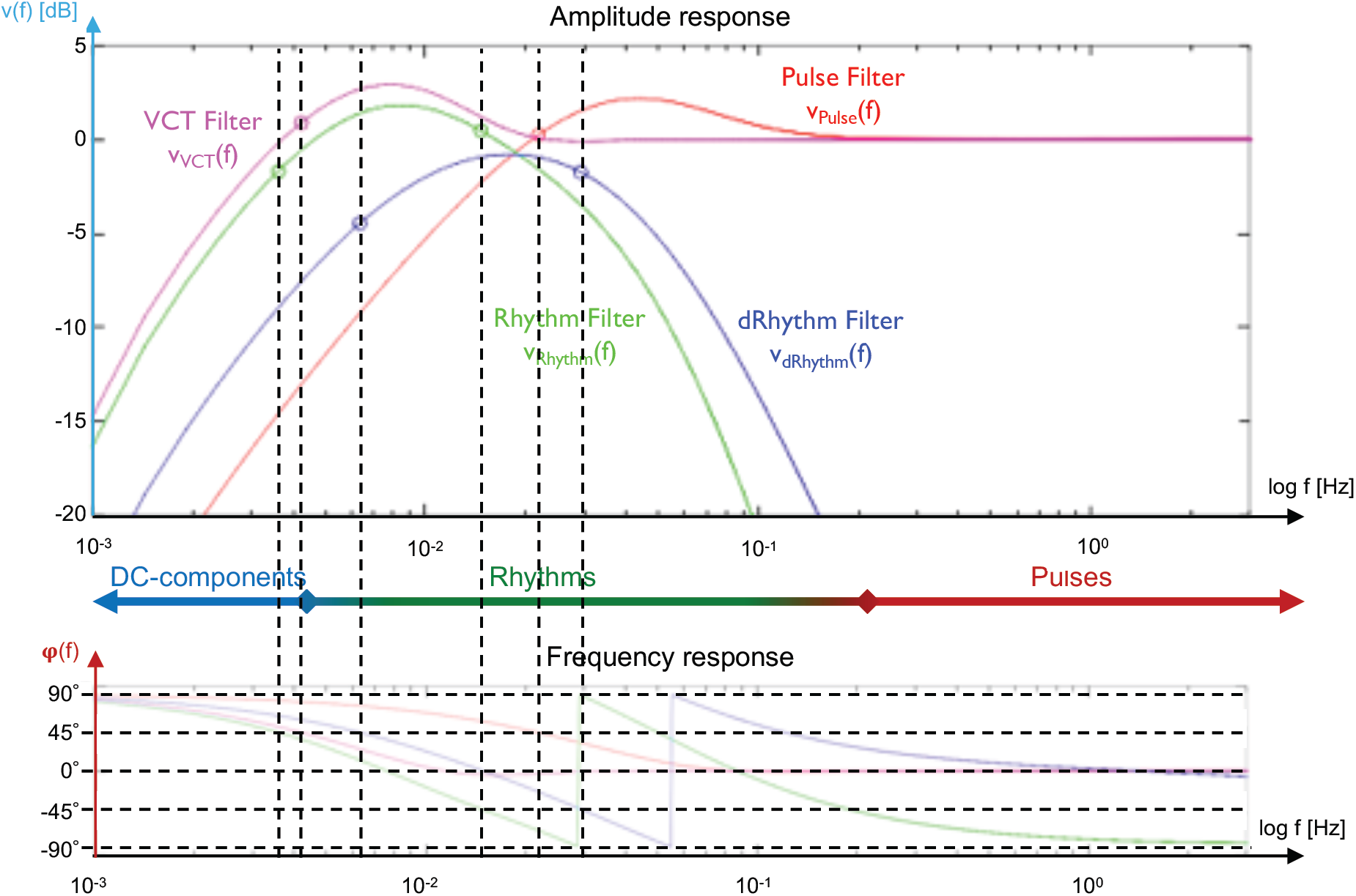
The frequency domain of the control signals, indicating which part of the BP rhythms is represented by the control signals. See text for further explanation.

### Tracking physiological rhythms

VCT ensures a stable long-term tracking of mBP, corrects for vasomotor activity and reconstructs BP information. In addition, a mechanism for tracking fast mBP changes and physiologic BP rhythms is needed. Figure 5 shows v_Rhythm_(f), which covers physiological rhythms in BP that are related to breathing as well as to “Traube-Hering-Mayer waves” [^29, 30^]. The signal v_Rhythm_(t) is led into a kind of a “proportional-integral-differential” (PID) control mechanism in order to keep v_Rhythm_(t) as small as possible by altering p_c_(t). Note that a rise in BP increases blood volume in the finger and thus decreases v(t), whereas a fall in BP increases v(t). The PID-control coefficients provoke negative feedback: if BP rises, p_c_(t) rises too while blood volume in the finger (indicated by v_Rhythm_(t)) is kept constant.

### Tracking pronounced BP changes

The so-called “differential” part of the controller, v_dRhythm_(f), also takes care of pronounced BP changes (e.g. Valsalva maneuver, hyperventilation, blood loss, etc.). In order to follow the overall CNAP2GO principle, where actuator changes shall be slow, pulsatile blood volume is not clamped. Rather, the pulsatile frequencies are cut off, and the “differential” controller part acts as a second band-pass filter producing non-pulsatile v_dRhythm_(f).

### Anti-resonance

The complex control system outlined above follows the inflection point and thus mBP is tracked by p_C_(t). As a result, the speed of tracking is limited to the cut-off frequency of v_Rhythm_(f) and v_dRhythm_(f). This produces the control engineering problem that the stability of the system is reduced if the actuator is slow. Slowly-reacting p_C_(t) creates a so-called pole in the z-plane indicating a tendency to oscillate. These oscillations especially occur when the patient calms down, parasympathetic tone increases, and the breathing frequency (about 0.2Hz to 0.3Hz) appears in the BP signal. A digital anti-resonance system was developed that makes this pole ineffective. The basic considerations of this system were inspired by a method for suppressing mechanical resonance in high track density hard-disk drives [^31^].

From CNAP2GO’s need for anti-resonance we see that a major benefit of pulsatile VUT control systems is to inhibit this kind of resonance: Even in very fast pressure generators, pressure in the finger cuff will always lag behind true pulsatile BP. However, the resulting pole in a VUT control system is ineffective as long as the system can be faster than the BP signal. Our anti-resonance system inhibiting the resonances also for non-pulsatile control is thus a major leap forward in the development of miniaturized BP measurement devices with acceptable clinical accuracy.

### Calibration

CNAP2GO’s p_C_(t) tracks mBP and, with a simple calculation, the pulsatile nature of blood pressure can be superimposed using v_Pulse_(t):

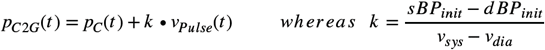

sBP_init_ and dBP_init_ are calibration values obtained before starting CNAP2GO and v_sys_ and v_dia_ are maxima and minima of v_Pulse_(t) during the initial phase.

### CNAP2GO prototype implementation

We implemented the CNAP2GO mechanism as a software prototype into existing hardware. A commercially available CNAP Monitor HD was adapted via software (see Methods section) to turn its VUT into CNAP2GO. Although the CNAP hardware in principle allows for the fast pulsatile BP changes as necessary for VUT, the speed of pressure changes was inherently limited by the software design of CNAP2GO as described before.

There were three reasons for this approach: First, the hardware of the CNAP Monitor HD has international regulatory approval for the use in patients. Second, the experiments could be performed prior to expensive investments into miniaturization. And finally, the CNAP Monitor HD allows calculation of all hemodynamic variables derived from the BP waveform.

CNAP2GO was implemented as software code (C++) and loaded as firmware on the CNAP Monitor HD. The combined system was granted a CE-mark for the use on patients in the OR. The sphygmomanometer of the CNAP Monitor HD was used for calibration.

### Lab tests versus CNAP Monitor HD

For obtaining CE-mark, lab tests in 20 healthy subjects were performed (Table 1). In Figure 6a, the BP and CO trends of the standard CNAP Monitor HD (VUT) and the CNAP2GO are shown. Both devices were initially calibrated to the same sBP/dBP values using the sphygmomanometer of the CNAP Monitor HD. In order to provoke the cardiovascular system and induce changes in BP and vasomotor activity, the subjects performed several physiological maneuvers (deep breathing, fast breathing, submersion of contralateral hand into ice water, stroop test, passive leg raining) within 30 min.

**Table 1:** Lab tests versus CNAP Monitor HD. Patient characteristics and results of Bland Altman analysis for mean arterial pressure. sBP – systolic blood pressure, dBP – diastolic blood pressure, mBP – mean arterial pres-sure, SD – standard deviation

|  |  |
| --- | --- |
| Patient characteristics |  |
| Male/female, n | 14/6 |
| Age, median (range) [years] | 36 (23–49) |
| Actual body weight, median (range) [kg] | 79 (56–130) |
| Height, median (range) [cm] | 177 (161–189) |
| CNAP Monitor HD sBP, median (range) [mmHg] | 108 (74–141) |
| CNAP Monitor HD dBP, median (range) [mmHg] | 64 (47–102) |
| CNAP Monitor HD mBP, median (range) [mmHg] | 82 (63–116) |
| Bland Altman analysis for mBP |  |
| Mean of the differences ± SD [mmHg] | 0.3 ± 4.4 |
| 95% limits of agreement [mmHg] | -8.3 to 8.9 |

**Fig. 6:**
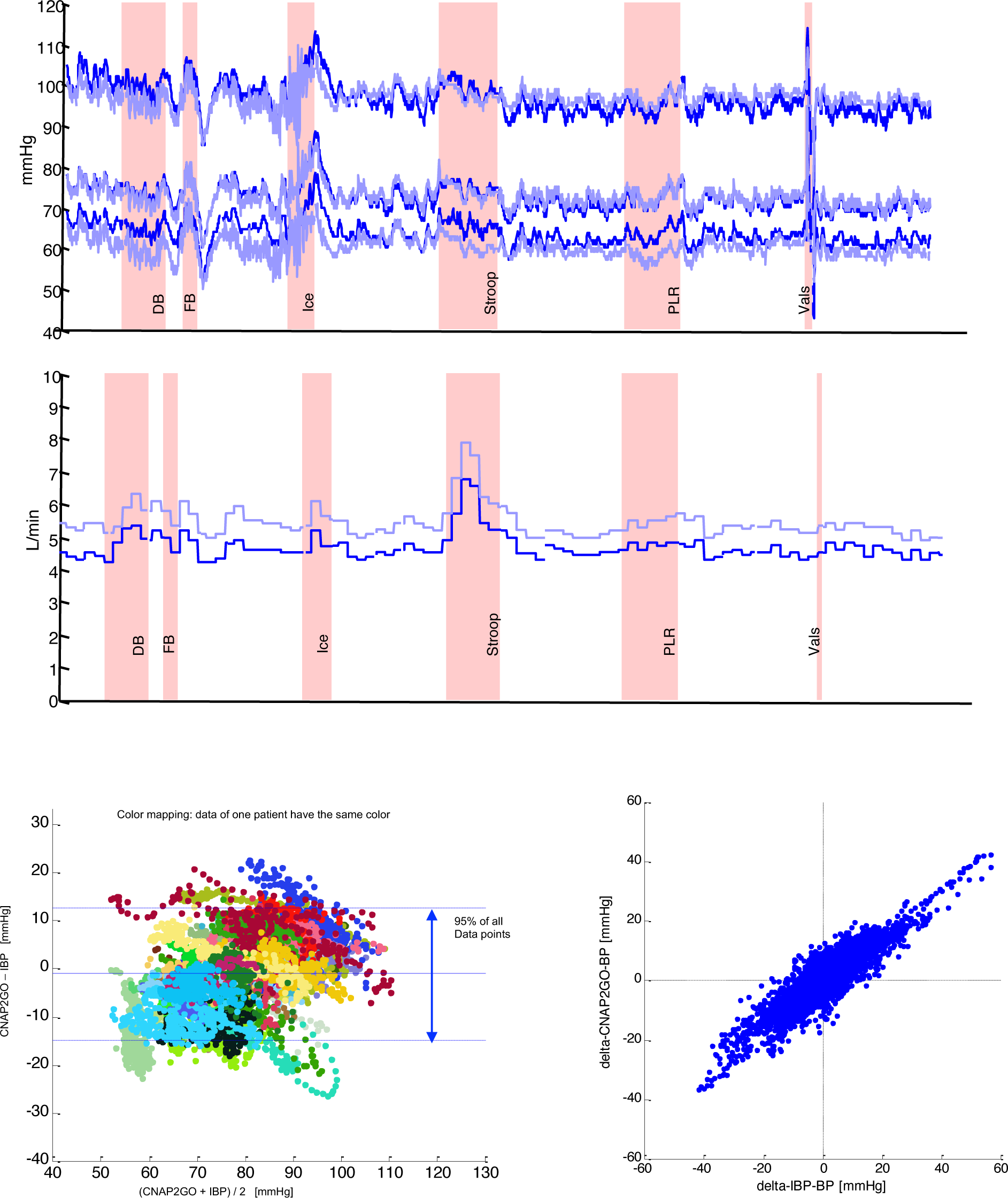
(a) Sample BP and CO trend traces taken from lab test measurements during physiological activities: DB – deep breathing, FB – fast breathing, Ice – submersion of hand in ice water, Stroop – colour stroop test, PLR – passive leg raising, Vals – valsalva test. The upper trace shows beat-to-beat sBP, dBP and mBP values derived by CNAP Monitor HD (dark blue) and by CNAP2GO (light blue). The lower trace shows CO derived by CNAP Monitor HD (dark blue) and by CNAP2GO (light blue). (b) Left panel: Bland Altman plot for mean BP from the clinical study: same-color points stem from the same patient. The plot includes indication of bias together with limits of agreement which define the range in which 95% of all data points are expected to lie. Right panel: Concordance plot of spontaneous changes in mBP found within 5 mins by invasive reference (IBP) and CNAP2GO.

The difference between mBP measured with CNAP2GO and mBP measured with CNAP Monitor HD was 0.3 ± 4.4 mmHg. The bias between CNAP2GO CO and CNAP Monitor HD CO was 1.4 ± 0.6 L/min with a percentage error of 22%. As shown in the sample trace of Figure 6a, there was an inherent parallel shift in the CO trend. This may be caused by a different shape of CNAP2GO’s BP pulse which is directly derived from the superimposed PPG-signal. The percentage error of 22% is below the 30% threshold that was defined by Critchley and Critchley to define clinically acceptable agreement between CO measurement devices [^32^].

### Clinical study versus invasive reference method

We included 46 patients having neurosurgery with general anesthesia in a clinical study. We compared mBP measured using CNAP2GO and mBP measured using a radial arterial catheter (invasive reference method, clinical gold standard) (Table 2). In the Bland Altman plot (Fig. 6b), we used color coding to illustrate measurements from the same patient, thus visualizing patient-specific offsets. Clustering indicates that patient-specific offsets remained similar throughout the recording which, in turn, indicates that CNAP2GO BP measurements were stable over the measurement period.

**Table 2:** Clinical study versus invasive reference method. Patient characteristics and results of Bland Altman analysis for mean arterial pressure. sBP – systolic blood pressure, dBP – diastolic blood pressure, mBP – mean arterial pressure, SD – standard deviation

|  |  |
| --- | --- |
| Patient characteristics |  |
| Male/female, n | 19/27 |
| Age, median (range) [years] | 56 (25–79) |
| Actual body weight, median (range) [kg] | 77.5 (46–130) |
| Height, median (range) [cm] | 171.5 (157–192) |
| Invasive sBP, median (range) [mmHg] | 104 (60–172) |
| Invasive dBP, median (range) [mmHg] | 56 (29–103) |
| Invasive mBP, median (range) [mmHg] | 78 (48–131) |
| Bland Altman analysis for mBP |  |
| Mean of the differences ± SD [mmHg] | -1.0 ± 7.0 |
| 95% limits of agreement [mmHg] | -14.8 to 12.7 |

The rate of actuator pressure change, which according to the VCT follows mBP, was analyzed in detail. The absolute changes in mBP of consecutive heart beats (N=97.432) were inspected together with the corresponding pulse intervals and resulting need for actuator speed. The contact pressure of CNAP2GO required a median changing speed of about 1.4 mmHg per beat or 1.3 mmHg/sec, the maximum values were 24.4 mmHg per beat or 25 mmHg/sec.

Figure 7 shows the spectra of the contact pressure p_c_(t) in comparison with the frequency content of the whole pulsatile BP signal, demonstrating how much the control was slowed down for CNAP2GO.

**Fig. 7:**
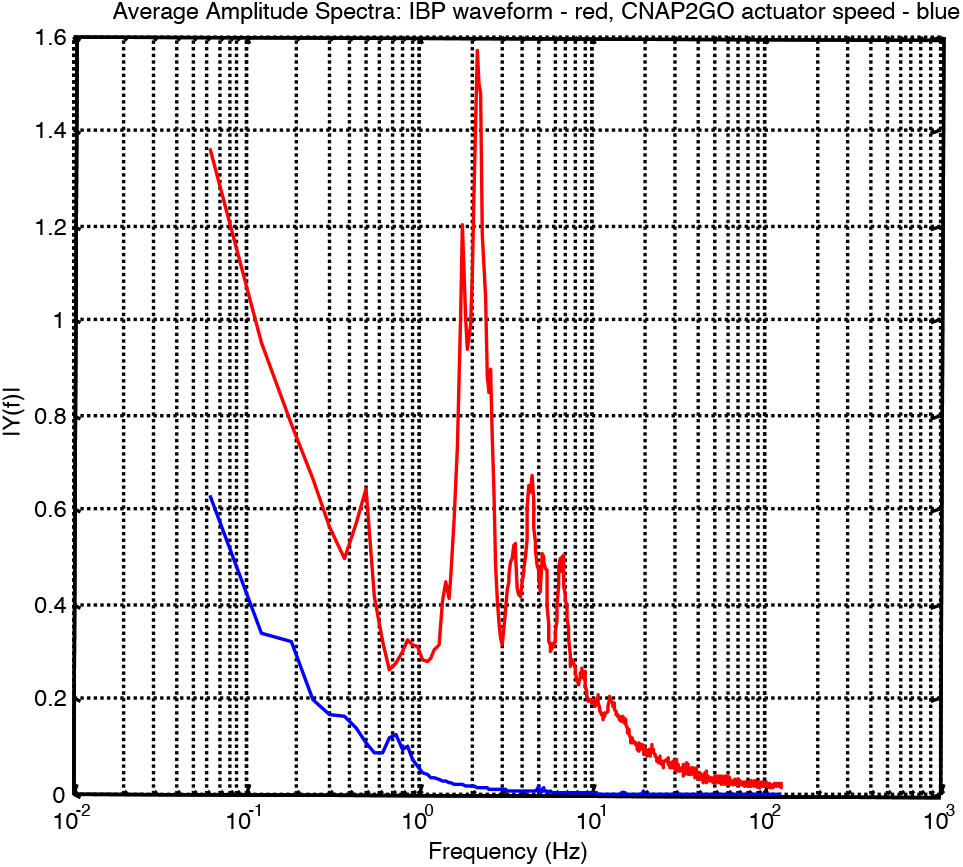
The spectra of CNAP2GO’s contact pressure p_c_(t) (blue) in comparison to the frequency content of the whole pulsatile BP signal (red).

### Important findings for the miniaturized actuator

The results of the clinical study reveal important findings for miniaturization of CNAP2GO. Miniaturized CNAP2GO requires an actuator with a maximum speed of at most 25-30 mmHg/sec. Actuators such as small printable piezo-electric motors are available with two important features: Low power consumption, but high enough stall torque to keep the actuator in its current position when no movement is needed. This has the effect that no energy is needed if mBP does not change.

We made basic calculations for a wearable CNAP2GO sensor using the data from a commercially available step motor (PCBMotor ApS, Ballerup, Denmark) to check whether the required pressure changes are feasible. We chose a printable motor with an outer diameter of 30 mm that can be integrated in the plate of a CNAP2GO finger ring. This motor can change contact pressure to the light elements via a gear from the motor. With a transmission ratio higher than 20.27, this component has a high enough stall torque so that no energy is needed if mBP does not change. With such a transmission system, the motor can theoretically change contact pressure of 1 mmHg within 5.82 ms. Assuming additional time due to control loops and further tolerances, we assume that a rate of pressure changes of 100 mmHg/sec can be easily achieved with this particular piezo motor.

In miniaturized CNAP2GO, readjustments of p_c_(t) to follow mBP will be done as soon as the beat detector of the system has found a beat (usually about 100ms after the systolic peak). Then the VCT criteria are used to derive the resulting new p_c_(t). In the previous experiments, p_c_(t) was adjusted using the CNAP pressure system and the finger cuff. In miniaturized CNAP2GO, the actuator will be activated until the new p_c_(t) is applied to the finger. Then the actuator stops and keeps p_c_(t) constant, at best, without power until the next beat is detected.

We have performed estimations on the power consumption of such a step motor. Assuming that it will follow the mBP profile of the 46 patients during anesthesia, the power needed to run the actuator is 44.83 mW. Together with the PPG system (5.64 mW), a low-power bluetooth microcontroller (1.66 mW) and a motion sensor (0.04 mW), the power requirement of the CNAP2GO system is less than 60 mW.

## Discussion

We have introduced the CNAP2GO method based on the new art of VCT which requires only slowly changing actuators and is robust against vasomotor changes. We discovered that the pulsatile control in the standard VUT mainly avoids control system resonances. For CNAP2GO, we achieved prevention of resonances by adaptive anti-resonance filters.

We were able to demonstrate validity of CNAP2GO. mBP differed by 1.0 ± 7.0 mmHg in comparison with the intra-arterial gold standard, which is well within the limits of 5 ± 8 mmHg demanded in the ISO 81060-2 standard for intermittent non-invasive sphygmomanometers [^33^].

There are still some challenges regarding the implementation of this method in a wearable solution: First, we didn’t perform our experiments using the final wearable hardware. Miniaturization will be a key task with this new sensor. Based on the present data set, all steps of miniaturization and technological refinement can be assessed and monitored in Lab-tests.

Second, for the wearable solution of CNAP2GO, the measured mBP needs a correcting element for orthostatic pressure whenever the finger level differs from heart level. Heart level correction can be done using a low-power motion sensor that measures the vertical difference between the heart and the finger. Knowing the density of blood, hydrostatic pressure can be calculated and simply subtracted from p_C_(t). This simple form of heart level correction has advantages compared to time-based wearable approaches, where the influence of this phenomenon is evident but solutions are unclear [^34^].

Third, for calibration, CNAP2GO must obtain initial absolute values of sBP and dBP. This may be achieved during the initial phase with the CNAP2GO sensor directly at the finger. Together with the initial mBP, a calibration function may be calculated from the measured oscillometric envelope and associated sBP and dBP.

In conclusion, we were able to demonstrate that the CNAP2GO method enables mBP to be measured with clinically acceptable accuracy and precision. Miniaturization of the hardware components can be realized using PPG sensors with an LED and a photodiode. The contact pressure of the light elements to the skin is modifiable by a slow and thus low-energy actuator, all of which make wearable CNAP2GO sensors feasible. CNAP2GO potentially constitutes the breakthrough for wearable sensors for blood pressure and flow monitoring in both ambulatory and inhospital clinical settings.

## Data Availability

Not applicable

## Data availability

Data that support hypotheses, plots and other findings of this study are available from the corresponding author upon reasonable request.

## Acknowledgements

The study was financed by CNSystems Medizintechnik GmbH, Austria. A major part of the investment came from a crowdfunding initiative (https://www.lionrocket.com/cnsystems) and the authors thank all crowd investors.

## Methods

### Hardware

To obtain a CNAP2GO protoype, we modified a standard CNAP Monitor HD, especially its core unit (the CNAP module). The CNAP Module is an electronic system where almost all components are implemented as software on a 32-Bit digital signal processor (DSP; TMS320F2810, Texas Instruments, Dallas, USA). Attached to the CNAP module is a double finger sensor system for the alternating use on the index and middle finger. Each CNAP sensor contains a PPG system utilizing an LED (λ=890nm) as well as a “light-to-frequency” converter (TSL245R, ams, Unterpremstätten, Austria) for light detection that produces a digital pulse-width modulation signal. This signal goes directly to a timer input of the DSP producing the digital PPG (i.e. the time series v(t)) [^35^]. The output of the DSP controls two valves (separate inlet and outlet valves). Both valves are arranged like transistors of a CMOS gate having one valve always closed. This allows for a high-fidelity contact pressure of the PPG-elements in the finger sensor [^36^].

### Algorithm overview

The original CNAP Monitor HD software in the module was replaced by the CNAP2GO/VCT method described in this article. All supporting functions (such as basic operating system, ambient light removal, valve controlling system, beat detector, data transmission, etc.) remained the same as in the original CNAP Monitor HD software V5.2.14.

### Initialization: Open-loop phase

In the open-loop phase, a Gaussian-style oscillometric envelope is calculated by using the amplitudes of v_Pulse_(t) obtained at different contact pressures p_c_(t). The set-point p_0_ and its PPG-counterpart v_0_ are found according to the maximum amplitude rule. The filter variables of the cascades v_Filt_[1, 2, ‥, N] are then set to v_0_.

### Filter cascade

After ambient light removal, v(t) has a sampling frequency (f_s_) of 250 Hz. A recursive digital IIR-filter was designed to obtain v_Pulse_(t), v_Rhythm_(t)), v_dRhythm_(t) and v_VCT_(t). Each filter cascade contains a first order IIR low-pass filter (y_i_ = (1-UC)*y_i-1_ + UC*x_i_). The recursive function provides down-sampling functionality to enable reasonable filter parameters for low cut-off frequencies. In Figure 5, the frequency response of the filters is shown.

### Closed-loop phase

After initialization in the open-loop phase, the filter adapts the relevant v-signals with every new data point. Signals “forget” the initial set-point v_0_ with an update coefficient UC [1, 2, ‥, N]. This corresponds to the fact that very low-frequency changes may be influenced by vasomotor activity. The information for low frequency BP changes is reconstructed by VCT [^37^, ^38^].

### Beat-based volume control technique

Within the VCT algorithm pulses are inspected whether they have become “spiky” or “fat” within the previous beat by calculating an integral. If the area over the negative half-wave differs from that over the positive half-wave, the integral over the complete beat is different from zero. This then triggers an adjustment of the set-point P_n_:

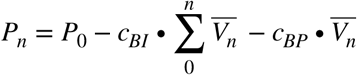

where *c*_*BI*_ and *c*_*BP*_ are constants for the beat-based integral (BI) and beat-based proportional (BP) control approach, respectively. The continuous summation starting with the first beat allows for the reconstruction of the long-term BP information that is filtered away from v_VCT_(t).

The integral function for the beat is:

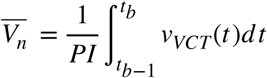

*t*_*b*_: time t at which beat b is detected and

PI: the pulse interval from *t*_*b*−1_ to *t*_*b*_

### Continuous control mechanisms

The CNAP2GO control system continuously keeps track of the required contact pressure with the following control structure:

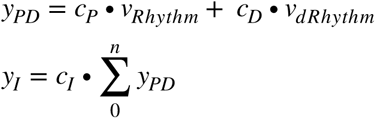

The final CNAP2GO contact pressure is:

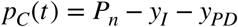

*c*_*P*_, *c*_*D*_ and *c*_*I*_ are constants for the proportional, differential, and integral approach of the continuous control system, respectively, that keeps p_c_(t) at the inflection point of the S-curve and thus makes it equal to mBP.

### The anti-resonance system

The resonance detector indicates frequency and amplitude of a resonance oscillation in p_c_(t), which typically occurs between 0.1Hz to 1Hz. This information tunes a high-sensitive IIR notch filter which eliminates resonance phenomena.

### Lab tests versus CNAP Monitor HD

Written informed consent was obtained from all participants. We performed measurements with the participants lying in supine position. The finger sensor of the CNAP2GO system was attached to the same hand as the CNAP Monitor HD sensor. Study measurements were performed over 30 min and included 6 physiological maneuvers.

For statistical analysis, simultaneously obtained mBP values measured with the CNAP2GO and with the CNAP Monitor HD were averaged over 10 sec to reduce the number of comparisons and de-correlate consecutive data. We performed Bland-Altman analysis accounting for multiple measurements per patient [^39^] and calculated the bias (mean of the differences between CNAP2GO mBP values minus CNAP Monitor HD mBP values), the standard deviation of the differences, and the 95% limits of agreement (Table 1).

### Clinical study versus invasive reference

The clinical prospective method comparison study comparing mBP measured using CNAP2GO and mBP measured using a radial arterial catheter (invasive reference method, clinical gold standard) was performed in the Department of Anesthesiology, Center of Anesthesiology and Intensive Care Medicine, University Medical Center Hamburg-Eppendorf (Hamburg, Germany) after approval by the ethic’s committee (Ethikkomission der Ärztekammer Hamburg, Hamburg, Germany). We obtained written informed consent from all patients. Adult patients scheduled for neurosurgical procedures in whom continuous blood pressure monitoring with an invasive arterial catheter was planned, independently of the study, were eligible for study inclusion. Exclusion criteria were the presence of vascular abnormalities or anatomical deformities of the upper extremities or peripheral edema.

The finger sensor of the CNAP2GO system was attached to the hand opposite to the arterial catheter. CNAP2GO continuous noninvasive BP measurements and invasive reference measurements using a radial arterial catheter were recorded simultaneously over a period of 45 minutes. According to clinical standards, general anesthesia was maintained with remifentanyl and propofol. Norepinephrine was given whenever clinically indicated. For statistical analysis, mBP measured with CNAP2GO and mBP obtained using an radial arterial catheter were synchronised in time before being averaged over 10 secs to reduce the number of comparisons and de-correlate consecutive data. We plotted mBP measured with the CNAP2GO system against invasively measured mBP values in scatter plots for visual assessment of the distribution and relationship of the BP data. In order to evaluate the agreement between mBP measured with CNAP2GO and mBP obtained using an radial arterial catheter we used Bland-Altman analysis accounting for multiple measurements per patient [^39^] and calculated the bias (mean of the differences between CNAP2GO mBP values minus invasively assessed mBP values), the standard deviation of the differences, and the 95% limits of agreement (see Table 2 and Fig. 6b left panel). Furthermore, we calculated the changes in BP (delta-BP) occurring over a period of 5 mins and investigated the concordance between changes in CNAP2GO mBP and changes in invasive reference mBP (see Fig. 6b, right panel).

